# Calculating the probability that a previously susceptible individual is infected as a function of time following exposure to SARS-CoV-2

**DOI:** 10.1101/2025.03.05.24317200

**Authors:** Alun Thomas, Karim Khader, Adam L Hersh, Matthew H Samore

**Affiliations:** Division of Epidemiology, Department of Internal Medicine, University of Utah School of Medicine, Salt Lake City, Utah, USA; Informatics, Decision-Enhancement, and Analytical Sciences Center of Innovation, VA Salt Lake City Health Care System, Salt Lake City, Utah, USA; Division of Infectious Diseases, Department of Pediatrics, University of Utah

**Keywords:** Isolation, quarantine, case counting, follow up testing

## Abstract

Reliable assessment of disease state probabilities for an individual following a specific exposure event, such as occupational exposure, is critical for managing isolation and quarantine to reduce exposure of further susceptible individuals to infection. We provide a method, programs, and web site for calculating the disease state probability of an individual immediately following an exposure to SARS-CoV-2 that may or may not have resulted in a transmission. We illustrate the utility of these computations by calculating the time at which an exposed individual’s risk of being infectious reaches an acceptable level, evaluating the benefit of a second test for a symptom free individual who has an initial negative test, assessing the value of PCR and antigen testing for case counting, and calculating the time at which the risk of an infected individual being still infectious reaches a level comparable with general background risk. Our results make it clear that evaluation of test results should not be done naively: a test may be negative because a transmission has not occurred, because the test result is false negative, because the individual has passed quickly through the course of the infection, or because the individual is experiencing an unusually long latent phase. Clearly, each of these situations has different implications and the value of our software is in differentiating, accurately assessing and clearly displaying the probabilities for all these disease states.

## 1 Introduction

Quarantine refers to the precautionary sequestration of individuals who have been exposed to a communicable disease. The purpose is to prevent further transmission of an infectious agent in case the exposed individual becomes infectious. Quarantine is particularly important when pathogens are high threat and when infected individuals can be infectious while asymptomatic. The recommended timing of release from quarantine is a key component of public health policies.

Generally, it is not feasible to conduct randomized trials or non-experimental intervention studies to estimate the relative effectiveness of alternative policies for quarantine. However, it is increasingly recognized that mathematical models can play a valuable role to inform control. A quantitative approach is useful for identifying parameters that should be incorporated into decision-making and for integrating existing knowledge about the epidemiological characteristics of an infectious disease, such as the distribution of time from infection to infectiousness, for example, the latent period.

The devastating nature of the coronavirus disease *(COVID-19)* pandemic motivated an intensive debate about guidelines for duration of quarantine. Policies changed as the pandemic progressed, reflecting the development of new options for diagnosis, prevention, and treatment, as well as the evolution of the virus itself. A salient issue for policy-making was to consider the impact of quarantine on the workforce of healthcare personnel available to manage the surge in admissions of critically ill patients.

In this paper, we describe a novel modeling method to support policy-making around termination of quarantine following a point exposure. We have parameterized the model to correspond to the original Delta strain of severe acute respiratory syndrome coronavirus 2 *(SARS-CoV-2)* as well as the Omicron variant. However, the approach is broadly applicable for other pathogens that are appropriately represented by an *SEIR, or Susceptible-ExposedInfectious-Recovered* model, and which include an infectious asymptomatic or presymptomatic state.

In the Methods section, we first describe the COVID-19 disease progression model we have assumed, indicate sources in the literature for model parameter estimates, and outline the nature of the computations. We then give the justification for our method of computing diseases state probabilities. The implementation of our methods in the `R` (R Core Team 2015) statistical environment and details of the online availability of the source code is then presented. We also describe a `shiny` website that runs our programs and can be used by the reader to make calculations and adjust the parameter values used.

In the Results section we first consider the disease state probabilities for an individual following an exposure event with a 30% probability of transmission. We additionally show the effect on these probabilities of observing the symptomatic state of the individual and of both polymerase chain reaction *(PCR)* and antigen test results. We then consider the added value of a second test for an individual who remains symptom free following an initial negative test. Following this we consider the time it takes for the risk of infectiousness of an infected individual to revert to that of a general member of the population. Finally, we compares the value of PCR and antigen tests for case counting and otherwise inferring a transmission event.

Throughout this work we use *infected* to mean that an individual is either in one of the active states of the disease, and hence infectious, or in the latent state and shortly to become infectious.

## 2 Methods

### 2.1 Disease progression model

Figure 1 gives a schematic of the possible paths we that model through a course of infection following an exposure to SARS-CoV-2. With probability `p`, the exposure results in an infection and the individual enters the latent state and progresses through subsequent phases. With probability 1 − `p` the individual remains susceptible until they become infected through general background exposure. Note that we distinguish two recovered states depending on whether or not the individual previously experienced symptoms. This is the sole distinction between these states and is introduced only as an artifice to simplify the computations described below. Probability parameters control the path taken, and the time in state variables are all assumed to be Gamma distributed.

**Figure 1:**
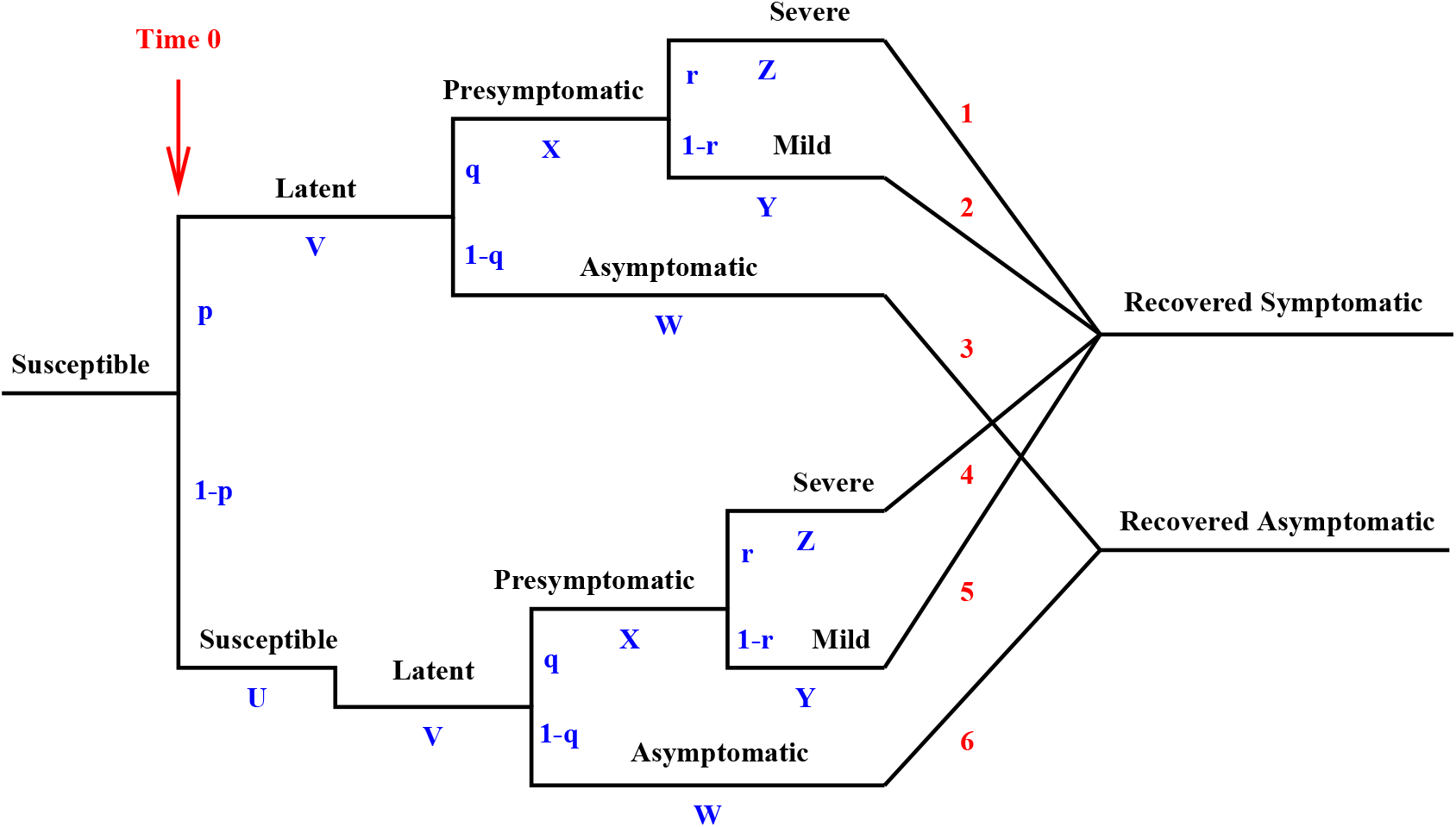
Model for the course of infection. Letters representing the random variables specifying the time in each state and branching probabilities are shown in blue. The red numbers enumerate the possible paths.

For the most part, to model the effects of the original variants of SARS-CoV-2, we chose values for the path splitting parameters and the Gamma shape and rate parameters to approximately match the values given in table 1 of Eikenberry et al. (2020). Tables 1 and 2 give details of these parameters. The columns marked *Lower* and *Upper* in table 1 give the lower and upper 2.5% quantiles of the these Gamma distribution and roughly correspond to the *Likely range* column given by Eikenberry et al. (2020). The values for the time in the susceptible state were arbitrarily chosen so that the background risk of infection is 1 per 1000 per day. The current daily risk in the US is approximately 1 per 10000 per day, but we use the higher rate to better reflect the risks to active individuals, allow for under reporting of cases, and model more intense stages of the epidemic. Note that the model used by Eikenberry et al. (2020) does not have a *presymptomatic* phase, so we assumed that this had a mean of 3 days with likely range of (1.0, 5.0) and reduced the times in the symptomatic phases accordingly.

**Table 1:** Gamma time-in-state distribution parameters and summary values for the original SARS-CoV-2 Delta variants.

| Parameter | Name | Shape | Rate | Mean | Variance | Lower | Upper |
| --- | --- | --- | --- | --- | --- | --- | --- |
| U | Sus | 1.00 | 0.001 | 1000.00 | 1000000.00 | 25.32 | 3688.88 |
| V | Lat | 3.47 | 0.680 | 5.10 | 7.50 | 1.22 | 11.70 |
| W | Asy | 6.53 | 0.933 | 7.00 | 7.50 | 2.71 | 13.30 |
| X | Pre | 9.00 | 3.000 | 3.00 | 1.00 | 1.37 | 5.25 |
| Y | Mld | 0.55 | 0.110 | 5.00 | 45.50 | 0.01 | 24.06 |
| Z | Svr | 2.66 | 0.242 | 11.00 | 45.50 | 1.97 | 27.62 |

**Table 2:** Probability parameter values specifying the path through the course of infection for the original SARS-CoV-2 variants.

| Parameter | Name | Value |
| --- | --- | --- |
| q | Symptomatic | 0.50 |
| r | Severe | 0.10 |

With regard to the later Omicron SARS-CoV-2 variants, Jansen et al. (2021) estimate that the latent period is roughly 60% that for the original variants. Symptomatic periods are estimated to be reduced by a factor of 90% (Menni et al. 2022), and we have assumed that both the presymptomatic and asymptomatic periods are similarly reduced. These changes have been made by reducing both the means and standard deviations of the Gamma distributions by the appropriate factor. Table 3 gives details of these Gamma distributions. The path probability parameters have been left unchanged.

**Table 3:** Gamma time-in-state distribution parameters and summary values for more recent SARS-CoV-2 Omicron variants.

| Parameter | Name | Shape | Rate | Mean | Variance | Lower | Upper |
| --- | --- | --- | --- | --- | --- | --- | --- |
| U | Sus | 1.00 | 0.001 | 1000.00 | 1000000.00 | 25.32 | 3688.88 |
| V | Lat | 3.47 | 1.133 | 3.06 | 2.70 | 0.73 | 7.02 |
| W | Asy | 6.53 | 1.037 | 6.30 | 6.08 | 2.43 | 11.97 |
| X | Pre | 9.00 | 3.333 | 2.70 | 0.81 | 1.23 | 4.73 |
| Y | Mld | 0.55 | 0.122 | 4.50 | 36.86 | 0.01 | 21.66 |
| Z | Svr | 2.66 | 0.269 | 9.90 | 36.86 | 1.78 | 24.86 |

Since we are primarily concerned with modeling the short term following exposure, we do not model a transition from the recovered state back to susceptible.

### 2.2 Probabilities of clinical observations and test results given underlying state probabilities

Observations about the individual following the exposure event will change the state probabilities. To account for these effects we need only specify the probabilities of such observations for each disease state, and then combine these with the above disease progression model using Bayes rule, as described below.

Regarding clinical observations, if an individuals is observed not to have shown any symptoms up to a particular time, the mild and severe disease states are excluded as is the recovered symptomatic state.

For test result data, we need to account for both qualitative and quantitative differences in PCR and antigen tests and there is also evidence that the performance of both types of test depend on viral load and can differ depending on the underlying state. For instance, it is likely that tests are more sensitive for severely symptomatic individuals than mildly symptomatic or asymptomatic individuals. The conventional two parameter specification of test performance in terms of sensitivity and specificity is, therefore, inadequate to convey the complexity we need. Instead we require a vector of values giving the probability of a positive test in each of the 8 disease states of our model. We label these *s*_*S*_, *s*_*L*_, *s*_*A*_, *s*_*P*_, *s*_*M*_, *s*_*V*_, *s*_*RS*_, *s*_*RA*_ respectively. Our model does not allow for test sensitivity to vary with time within any state.

Studies consistently show that both test specificities are very high, for instance, the large meta analysis of Dinnes et al. (2022) estimates specificity of antigen test to be above 99.5% over a broad rage of brands. We, therefore, set *s*_*S*_, the probability of a true negative testing positive, at 0.5% for both antigen and PCR tests. Similarly, we have *s*_*L*_ = 0.5%.

Liu et al. (2022), based on agreement between multiple independent tests on over 3500 subjects, estimated the sensitivity of PCR tests to be greater than 95% with no negative results for confirmed positives until late in the recovery phase. Smith-Jeffcoat et al. (2024) had a lower estimate with sensitivity peaking at 83% 3 days after onset of symptoms or 3 days after an initial positive test for asymptomatic individuals. Their results also suggests a lower sensitivity of around 20% before the symptomatic phase, however, this includes not only the presymptomatic phase for symptomatic people but also the latent phase for those who will be asymptomatic. This study was smaller than that of Liu et al. (2022), and the dependence of sensitivity on how subjects were referred to the study suggests that some of their results are more to do with the true status of their subjects rather than on intrinsic test properties. For PCR tests, we, therefore, follow the more favourable sensitivity estimates and set *s*_*A*_, *s*_*P*_, *s*_*M*_, *s*_*V*_, *s*_*RA*_, and *s*_*RS*_ at 95%.

Both Dinnes et al. (2022) and Smith-Jeffcoat et al. (2024) find considerable variation in antigen test sensitivities across studies by brand of test and by phases of infection. Overall, Smith-Jeffcoat et al. (2024) estimate antigen sensitivity at just below 50% that of PCR tests. Dinnes et al. (2022) give somewhat higher estimates. Both find substantially higher sensitivity among symptomatic (65%-73%) compared with asymptomatic individuals (20%-55%), and Smith-Jeffcoat et al. (2024) noted even higher sensitivity (80%) among individuals with fever. Both studies showed sensitivity decreasing strongly after the first week of infection. While we are not able to accommodate all these findings in our model for antigen tests, setting *s*_*A*_ = 50%, *s*_*M*_ = 70% and *s*_*V*_ = 80% is broadly in line. As with PCR tests Smith-Jeffcoat et al. (2024) shows lower sensitivity early on, but again this covers both the latent and presymptomatic states in our model so we set *s*_*P*_ = 50%. In contrast, they show that while PCR tests maintain sensitivity into the recovery phase, antigen tests sensitivity decreases to near the baseline within 1 week for asymptomatic people and within about 2 weeks for symptomatic, whether with or without fever. Hence, we set *s*_*RS*_ = *s*_*RA*_ = 0.5%.

There is no compelling evidence in the literature suggesting different test performance for different SARS-CoV-2 strains.

Table 4 gives the test parameters that we assume based on the above considerations for PCR and antigen tests for each of the underlying disease states. The relevant effects of these observational models are summarized, using the probability notation we define in section 2.3, in table 6.

**Table 4:**
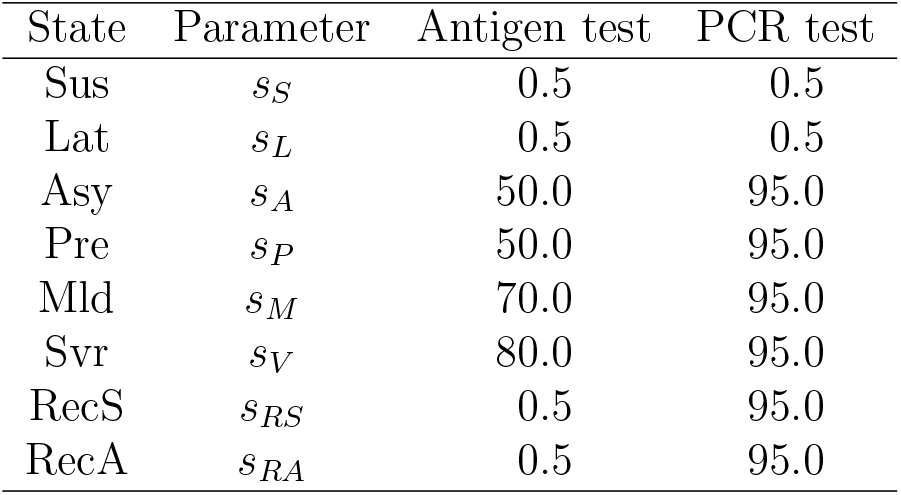
Assumed probabilities of a positive test, given as percentages, by test type and underlying disease states.

| State | Parameter | Antigen test | PCR test |
| --- | --- | --- | --- |
| Sus | $s_S$ | 0.5 | 0.5 |
| Lat | $s_L$ | 0.5 | 0.5 |
| Asy | $s_A$ | 50.0 | 95.0 |
| Pre | $s_P$ | 50.0 | 95.0 |
| Mld | $s_M$ | 70.0 | 95.0 |
| Svr | $s_V$ | 80.0 | 95.0 |
| RecS | $s_{RS}$ | 0.5 | 95.0 |
| RecA | $s_{RA}$ | 0.5 | 95.0 |

### 2.3 Probability calculations

We calculate the state probabilities following an exposure event by conditioning on the paths taken through the course of the infection. The paths, states, variable and parameter names used here are those used in figure 1.

We let *H* ∈ {1, … 6} index the path and let *S*_*x*_ ∈ {Sus, Lat, Asy, Pre, Mld, Svr, RecS, RecA} be the state the individual is in at time *x* days following the exposure event. Table 5 gives *P* (*H*), the path probabilities derived from the splitting probabilities, and *P* (*S*_*x*_|*H*), the conditional probabilities of being in each state *S*_*x*_ given that path *H* is followed. We use *F*_?_ as shorthand for the distribution function of sums of arbitrary Gamma distributed random variables. Thus, for instance,

**Table 5:** The probability, *P* (*H*), of following each possible path *H* of progression through the infection, and the probability of being in each state at time *x* given the path, *P* (*S*_*x*_|*H*).

| | Path $H$ | | | | | |
| --- | --- | --- | --- | --- | --- | --- |
|  | 1 | 2 | 3 | 4 | 5 | 6 |
| | $P(H)$ | | | | | |
| | $pqr$ | $pq(1-r)$ | $p(1-q)$ | $(1-p)qr$ | $(1-p)q(1-r)$ | $(1-p)(1-q)$ |
| | $P(S_x\vert H)$ | | | | | |
| Sus | 0 | 0 | 0 | $1 - F_U$ | $1 - F_U$ | $1 - F_U$ |
| Lat | $1 - F_V$ | $1 - F_V$ | $1 - F_V$ | $F_U - F_{UV}$ | $F_U - F_{UV}$ | $F_U - F_{UV}$ |
| Asy | 0 | 0 | $F_V - F_{VW}$ | 0 | 0 | $F_{UV} - F_{UVW}$ |
| Pre | $F_V - F_{VX}$ | $F_V - F_{VX}$ | 0 | $F_{UV} - F_{UVX}$ | $F_{UV} - F_{UVX}$ | 0 |
| Mld | 0 | $F_{VX} - F_{VXY}$ | 0 | 0 | $F_{UVX} - F_{UVXY}$ | 0 |
| Svr | $F_{VX} - F_{VXZ}$ | 0 | 0 | $F_{UVX} - F_{UVXZ}$ | 0 | 0 |
| RecS | $F_{VXZ}$ | $F_{VXY}$ | 0 | $F_{UVXZ}$ | $F_{UVXY}$ | 0 |
| RecA | 0 | 0 | $F_{VW}$ | 0 | 0 | $F_{UVW}$ |

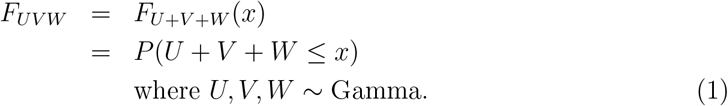

The state probabilities are then given by

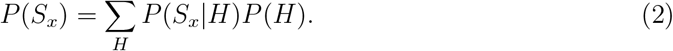

The specific path splitting probabilities and Gamma shape and rate parameters used here are given in tables 1, 2 and 3, but our implementation allows the user to change these values.

To calculate the conditional probabilities given in the results section, we consider the probability of some observation *O*_*x*_ at time *x* and specify *P* (*O*_*x*_|*S*_*x*_), the probability of the observation given the underlying state at that time. Then, using Bayes rule, we have

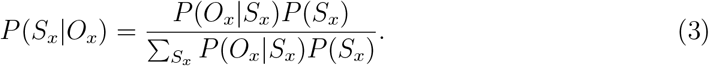

Table 6 specifies the conditional probabilities required for the following events:

**Table 6:** For each state we give *P* (*M*_*x*_ = 0|*S*_*x*_), the probability of no symptoms up to time *x* given the state, *P* (*I*_*x*_ = 1|*S*_*x*_), the probability of being infected given the state, *P* (*T*_*x*_ = 0|*S*_*x*_, Negative test), the probability of having a negative test given the state. The values in the final column will depend on the test, antigen or PCR.

| $S_x$ | $P(M_x = 0\vert S_x)$ | $P(I_x = 1\vert S_x)$ | $P(T_x = 0\vert S_x, \text{Negative test})$ |
| --- | --- | --- | --- |
| Sus | 1 | 0 | $1 - s_S$ |
| Lat | 1 | 1 | $1 - s_L$ |
| Asy | 1 | 1 | $1 - s_A$ |
| Pre | 1 | 1 | $1 - s_P$ |
| Mld | 0 | 1 | $1 - s_M$ |
| Svr | 0 | 1 | $1 - s_V$ |
| RecS | 0 | 0 | $1 - s_{RS}$ |
| RecA | 1 | 0 | $1 - s_{RA}$ |

- *I*_*x*_ indicates that the individual is uninfected (0), or infected (1), at time *x*,
- *M*_*x*_ indicates that the individuals has not (0) or has (1) exhibited symptoms up to time *x*,
- *T*_*x*_ indicates that a test at time *x* is negative (0) or positive (1).

We note that the states RecS and RecA differ only in the path taken to get to them and we use this convention only because it allows us to distinguish between resolved individuals who have or have not shown symptoms in the past without explicit reference to the path, thus simplifying the first column of table 6.

To address the inferential problem of whether an exposure at time 0 resulted in an infection we let *E*_0_ be and indicator for the event that an infection occurred (1) or didn’t (0). We can then calculate the required likelihoods given observations *O*_*x*_ as

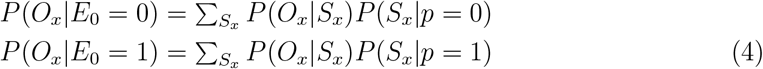

and if *P* (*E*_0_ = 1) is the prior probability that an infection occurred, the posterior given *O*_*x*_ is

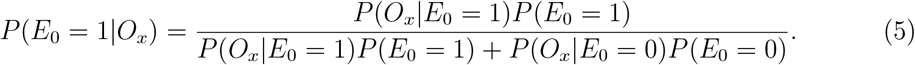

### 2.4 Implementation

We wrote programs to implement the above methods using the `R` statistical programming environment (R Core Team 2015). These are reasonably straightforward and short and are distributed in a single `R` source code file called `covid.R` which is available from `github` at

https://github.com/alun-thomas/covidisolation/ and, after downloading, can be loaded into an `R` environment using

> source(“covid.R”)

The programs are described in the document `covid.pdf` which is also available at the `github` site as is the file `isol.R` that provides the specific code necessary to produce the tables and figures in this paper which may be a useful example for those wishing to use or extend the methods described here. The files `covid.R`, `covid.pdf`, and `isol.R` are also provided as supplements to this article.

An interactive `shiny` application for calculating the state probabilities following an exposure using these programs is available at

https://alunthomas.shinyapps.io/covidisolation/

The application allows the user to input parameters and change the conditioning events. A screenshot of the application is shown in figure 2.

**Figure 2:**
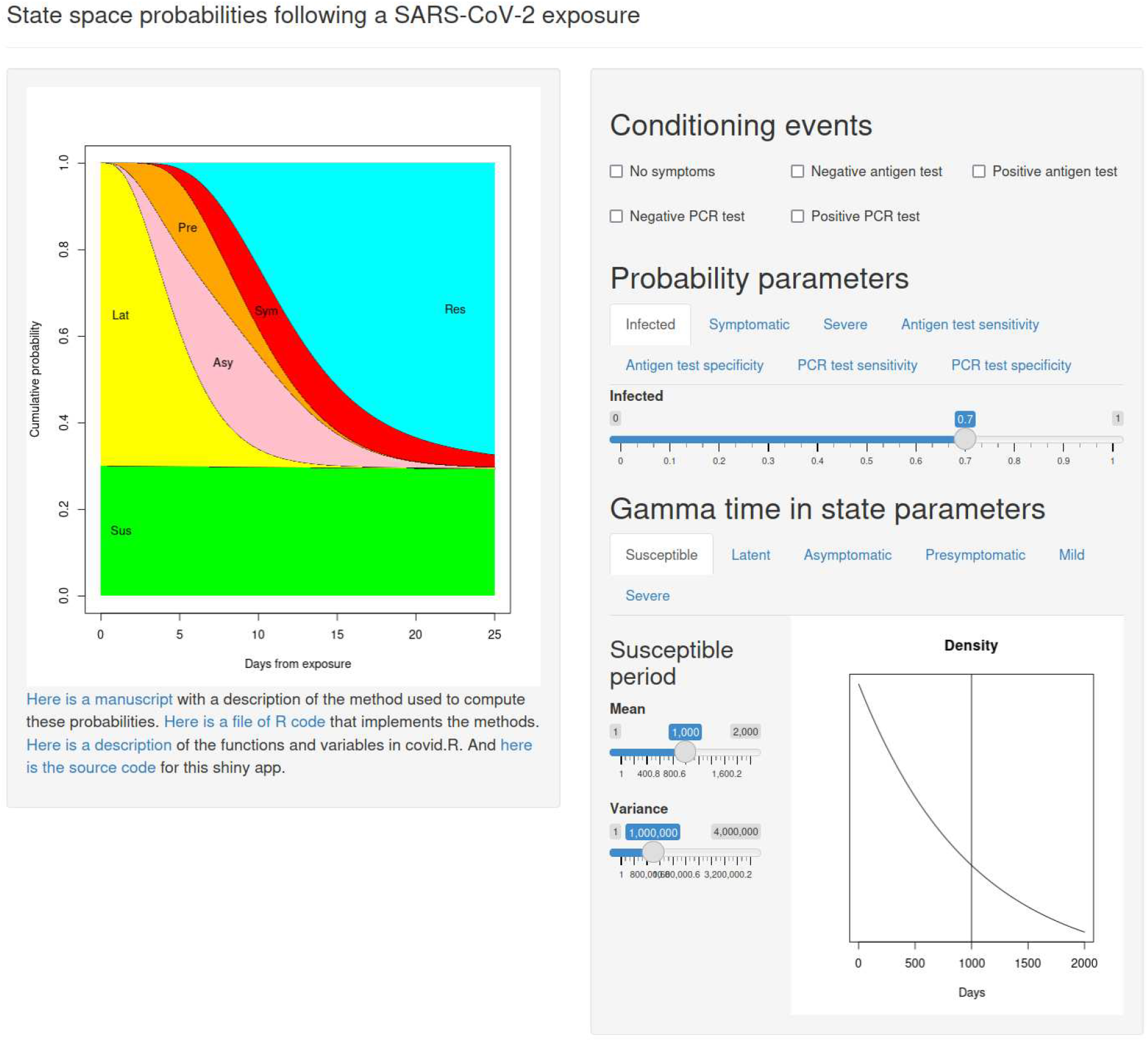
A screen shot of the `shiny` application for calculating SARS-CoV-2 state probabilities.

Our programs allow specifying arbitrary path splitting probabilities and arbitrary shape and rate parameters for the Gamma time in state variables. These calculations require the `pcoga()` function in the `coga R` package (Hu et al. 2021, Hu et al. 2020) to calculate the density functions of sums of independent Gamma random variables which can be installed using the standard `R` command

> install.packages(“coga”) and is also available at https://cran.r-project.org/web/packages/coga/index.html

Our source distribution includes programs to calculate these probabilities by simulating infection histories for individuals over a period of time following exposure. This was done in the straightforward way suggested by figure 1 using the standard `R` functions `runif()` to generate Uniform random variates for the path splitting events and `rgamma()` to generate the time-in-state values. We checked that the exact numerical and the simulation methods gave the same results within the sampling error of the simulation approach. The simulations are quick and the results are visually almost indistinguishable from the exact values when using 10000 simulated individual histories. More simulations may be needed if rejecting histories to condition on specific rare events.

## 3 Results

### 3.1 Disease state probabilities

First, we describe use of the tool to inform decisions regarding duration of quarantine. One approach for deciding when quarantine can end is to set a threshold probability for being infected, at or below which the person would be released from quarantine. In this context, being infected is defined as either being infectious or in the latent state, but does not include individuals who reached the recovered, non-infectious state.

For demonstration purposes, we illustrate the situation where exposure carried a 30% probability of transmission, contrasting two different thresholds, 10% and 1% which may represent different levels of acceptability to exclude infection depending on circumstances. For comparison purposes, the graphs are constructed to depict state probabilities following an exposure under four different monitoring policies: (a) neither symptom tracking nor testing; (b) symptom tracking alone; (c) symptom tracking plus antigen testing; (d) symptom tracking plus PCR testing. For the Delta variant, in the absence of symptom monitoring or testing, the 10% threshold and 1% thresholds were reached at 14.5 and 41.75 days, respectively (Figure 3). If an individual remained symptom free, the 10% and 1% thresholds were reached at 12.25 and 23 days, respectively. If the individual remained asymptomatic, a negative antigen test at days 10.0 and 19.75 corresponded to the 10% and 1% thresholds, respectively. If the individual remained asymptomatic, a negative PCR test at days 7 and 15 corresponded to the 10% and 1% thresholds, respectively.

**Figure 3:**
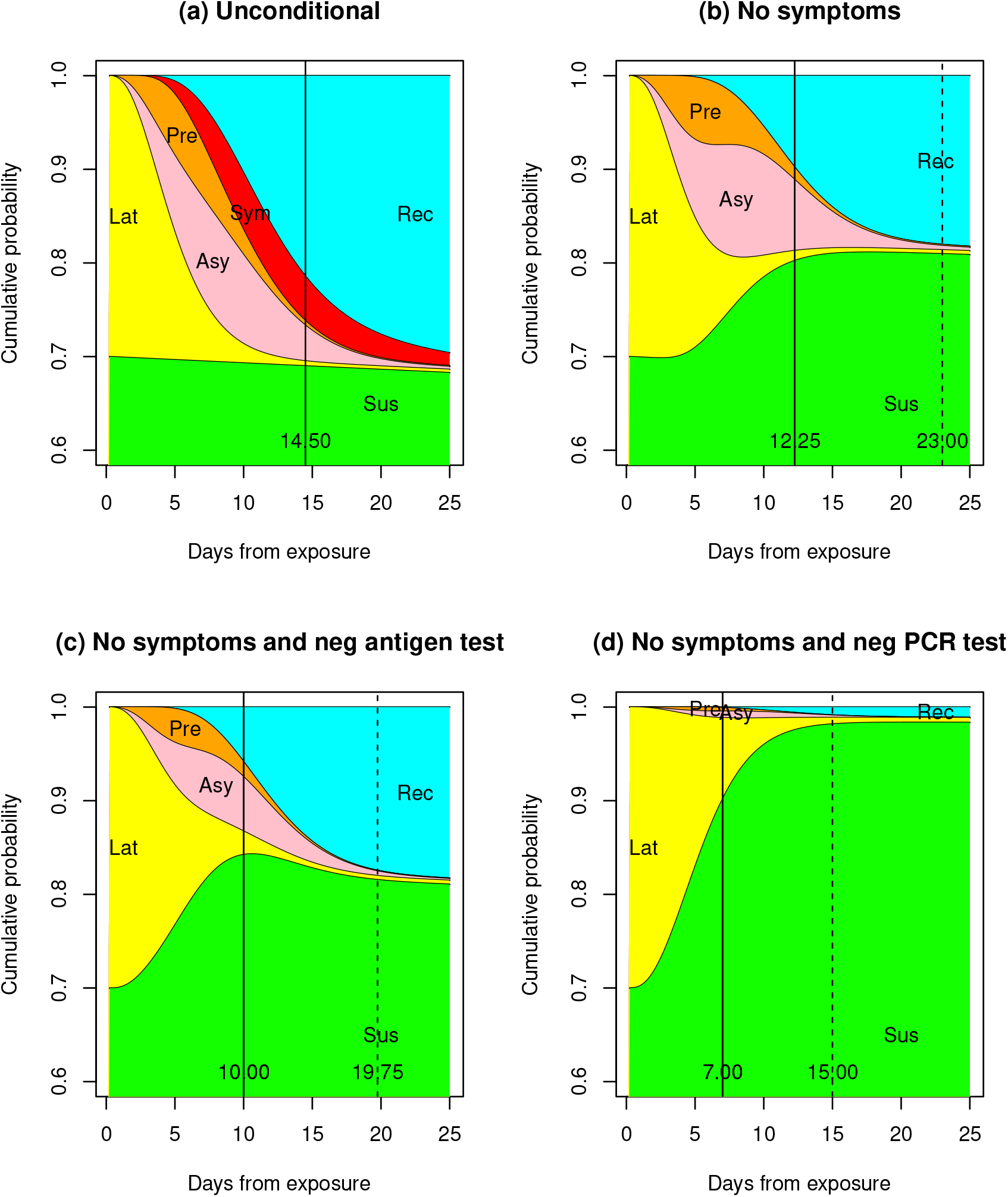
State probabilities following a 30% chance of transmission at exposure to the SARS-CoV-2 Delta variant. This shows the unconditional probabilities and those when no symptoms and negative test results are observed. The vertical black lines show where the probability of the infected states decreases to 10% (solid) and 1% (dashed).

When the same analysis was performed for the Omicron variant, the length of time to reach the 10% probability threshold was by about 3.5 days shorter; the amount of shortening across monitoring and testing scenarios ranged from 2.75-3.5 days (Figure 4). The length of time to reach the 1% threshold was 12 days shorter with no tracking, and between 3.0 and 7.5 days with tracking.

**Figure 4:**
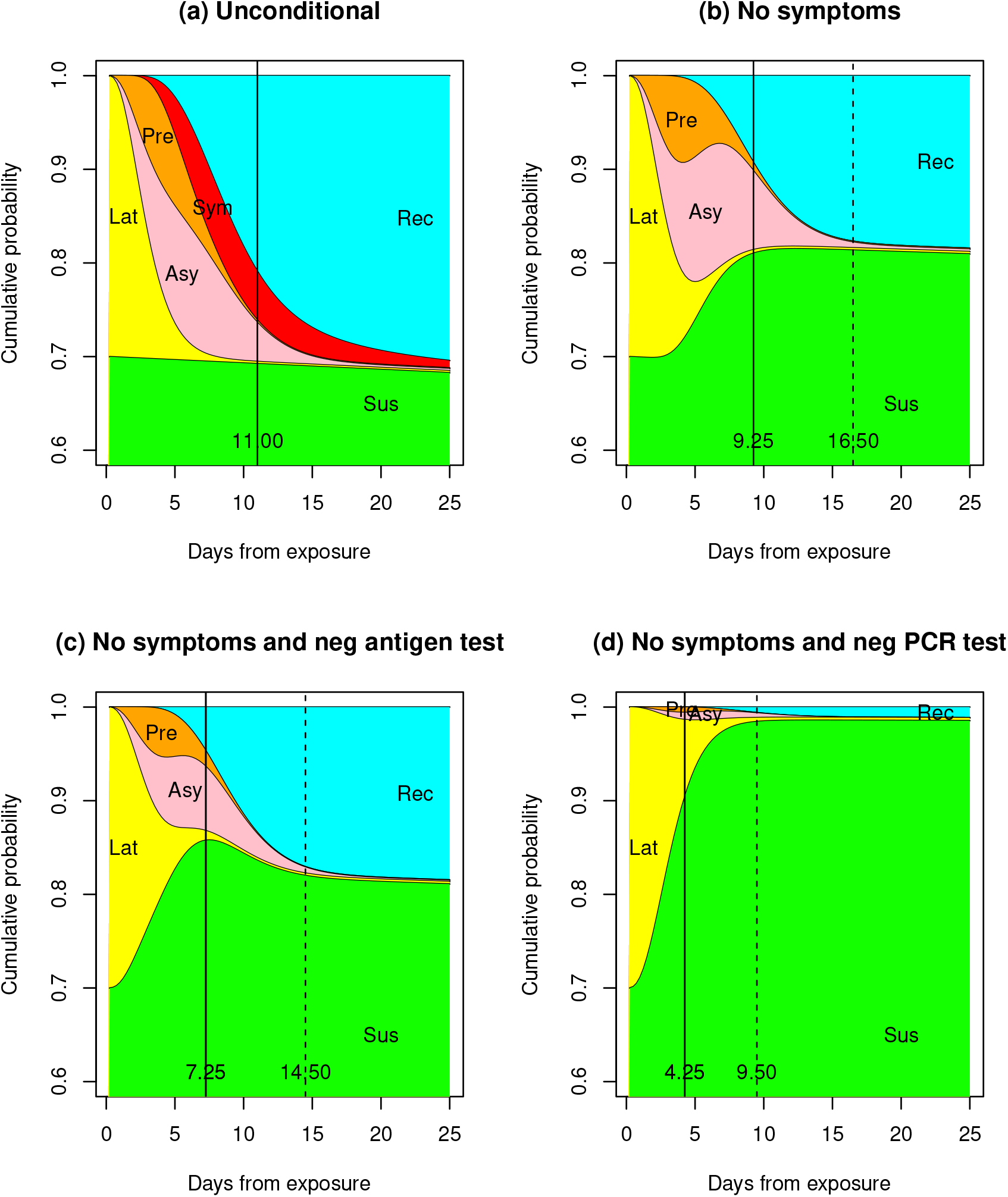
State probabilities following a 30% chance of transmission at exposure to the SARS-CoV-2 Omicron variant. This shows the unconditional probabilities and those when no symptoms and negative test results are observed. The vertical black lines show where the probability of the infected states decreases to 10% (solid) and 1% (dashed).

We note that the implications of a later negative test depends on whether it is antigen or PCR. For the antigen test, negativity can be evidence for there having been no initial transmission, or that the individual has passed through the infection to the recovered phase. A negative PCR test, on the other hand, is strong evidence for no transmission.

### 3.2 Follow up testing

We now describe use of our methods to assess the impact of performing a second test following a negative initial test. As before, the original exposure was presumed to carry a 30% risk of transmission, however, in this model, individuals were assumed to have already reached the 10% probability threshold based on absence of symptoms and timing of their initial negative test. Simulation was then used to determine the point in time when the probability of being in an infectious or latent state further decreased to 1%. In the scenario of symptom monitoring alone for the Delta variant, this threshold was not reached until day 24, (Figure 5). A negative PCR follow up test reduced this time to 19 days, and an antigen follow up to 14 days. These reductions were the same regardless of which test was initially carried out. For the Omicron variant, the pattern was similar with time to reaching 1% reducing from 16.5 days for a symptom free individual to 14 days with an additional negative antigen test and 8 days with an additional negative PCR test. Again, this pattern was only minimally affected by which test was initially used.

**Figure 5:**
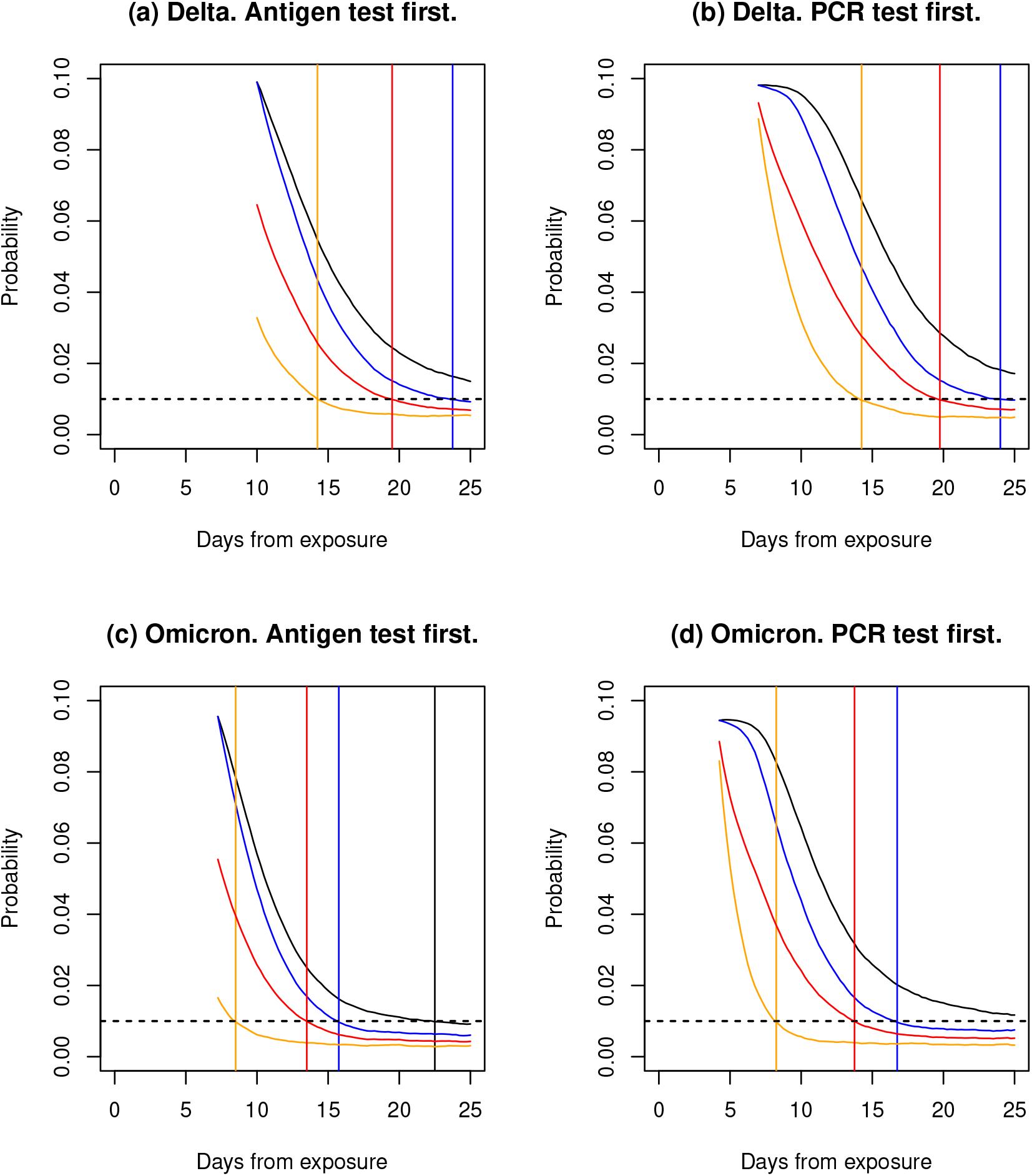
Plots of the probability that an individual is infected given that they are symptom free and have an initial negative test. These are presented for both Delta and Omicron variants and for the case when the initial test is an antigen test and a PCR test. The black lines are for individuals with no follow up, blue for individuals who are observed to remain symptom free, red for symptom free individuals with a further negative antigen test and orange for symptom free individuals with a further negative PCR test. The vertical lines indicate where the curves first cross the 1% threshold.

### 3.3 Effect of testing on assessing infectiousness

Here we consider the hypothetical situations where the probability of transmission following an exposure is either zero or one. For each of these boundary conditions, we plot the probabilities of being latent or infectious as a function of days following exposure. At the time that these curves cross, the risk posed by individuals who were certainly infected becomes equivalent to the general members of the population, as represented by the individuals who were certainly not infected from exposure. Within the assumptions of our model, the probability of infection among those who were certainly infected from exposure then drops below the background risk of infection for a period of time after the crossing point. The curves for the cases where we assume an intermediate probability of transmission will be numerical averages of the two extreme cases and, therefore, will share the same crossing points, which is why, for clarity of exposition, we consider only the extremes. Figure 6 shows plots for these probabilities, and also how they are affected by observing that an individual shows no symptoms and has a negative SARS-CoV-2 test.

**Figure 6:**
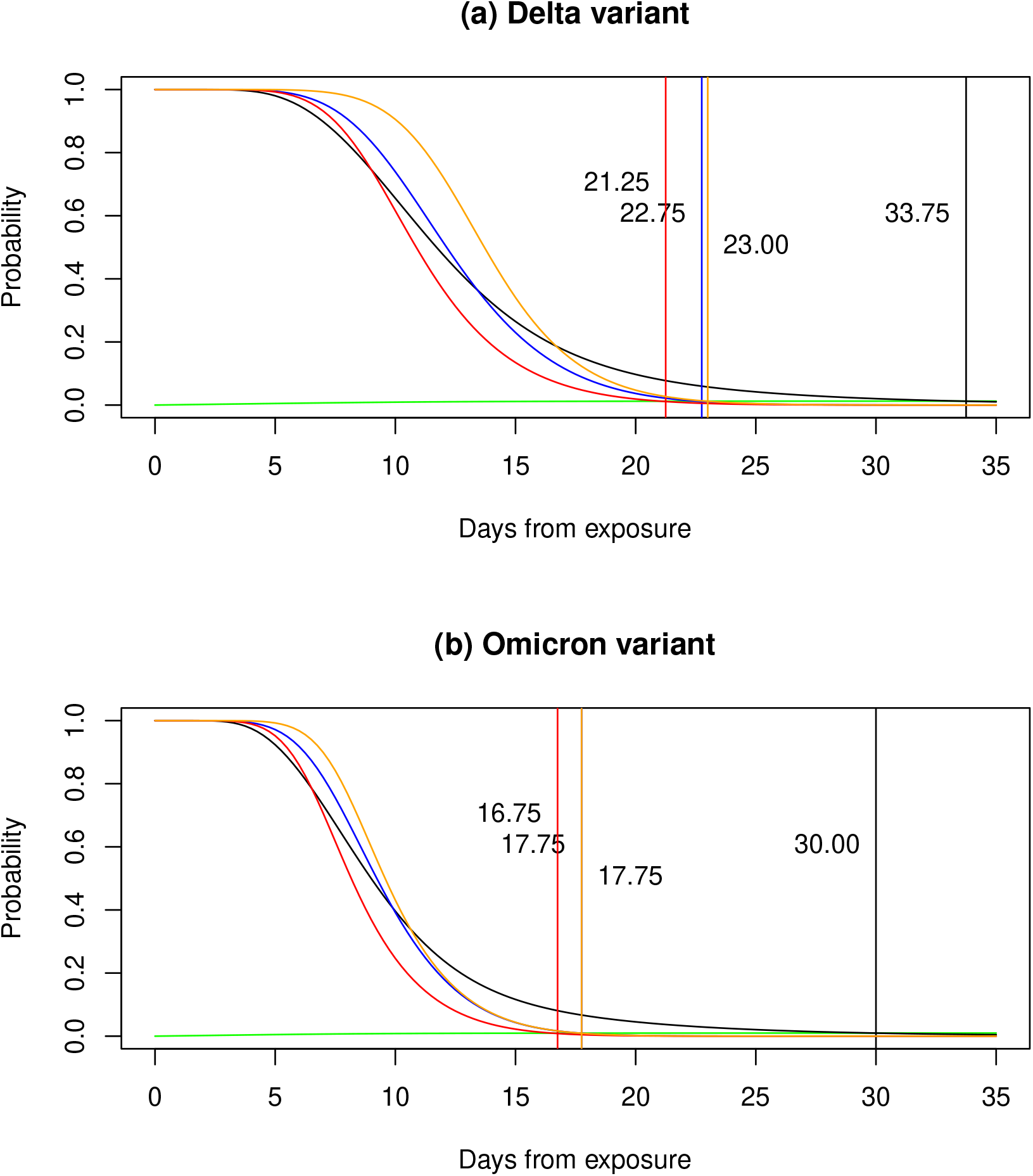
The probability that an individual who was certainly infected at time zero is still infectious at a given time following is shown in black. The blue line shows the probability for and individual who is asymptomatic up to the given time, the red line for in asymptomatic individual who has a negative Antigen test at the given time, the orange line for in asymptomatic individual who has a negative PCR test at the given time. The green line shows the probability for an individual who was certainly not infected at time zero, but was subsequently exposed to background risk of infection. Vertical lines in the appropriate colour show where the probabilities for certainly infected individuals first fall below those for certainly uninfected.

The black lines in figures 6(a) and (b) show that it takes on average almost 5 weeks, 33.75 days for Delta, 30 days for Omicron, for an individual to revert to presenting background levels of risk. Monitoring symptoms and testing can, however, detect those who have progressed quickly and, hence, present lower risk sooner, at a much earlier time. Symptom free individuals reach background risk at 22.75 days for Delta and 17.75 days for Omicron as shown by the blue lines Note that a negative PCR test does not give much additional information when no symptoms are observed, as shown by the closeness of the blue and orange vertical lines in figure 6(a). For the Omicron variant, figure 6(b), the vertical lines showing the crossing points are indistinguishable.

Conversely, a negative antigen test moves the crossing point 1.75 days earlier for Delta, and 1 days earlier for Omicron as shown by the red lines. This is because a negative PCR test can select for individuals with an unusually long latent period, while a negative antigen test detects those individuals who have progressed quickly through the whole course of the infection. Thus, for purposes of assessing the risk an infected individual poses to others, an antigen test is more somewhat more informative.

### 3.4 Inferring transmission events

We now consider the value of testing for determining whether transmission actually resulted from the exposure. Curves for a range of prior probabilities were plotted, the prior bing represented by the y-axis values at time 0. Bayesian analysis was used to calculate posterior probabilities as a function of time since exposure, comparing similar monitoring strategies as before (Figure 7). We again focused on negative evidence because the tests are highly specific and the consequences of positive results are clear. Under the assumption that the antigen test is negative during the recovered state, the antigen test adds comparatively little information compared to symptom monitoring alone, for the objective of inferring transmission (figures 7(c) and (d) versus 7(a) and (b)). In contrast, the negative predictive value of the PCR test is much higher, as denoted by the substantial reduction in probability of transmission when the PCR test is negative (figures 7(e) and (f)). Thus, for purposes of case counting, the PCR test is better. Performing the PCR test at least 10 days following exposure largely avoids the risk of testing during the latent period.

**Figure 7:**
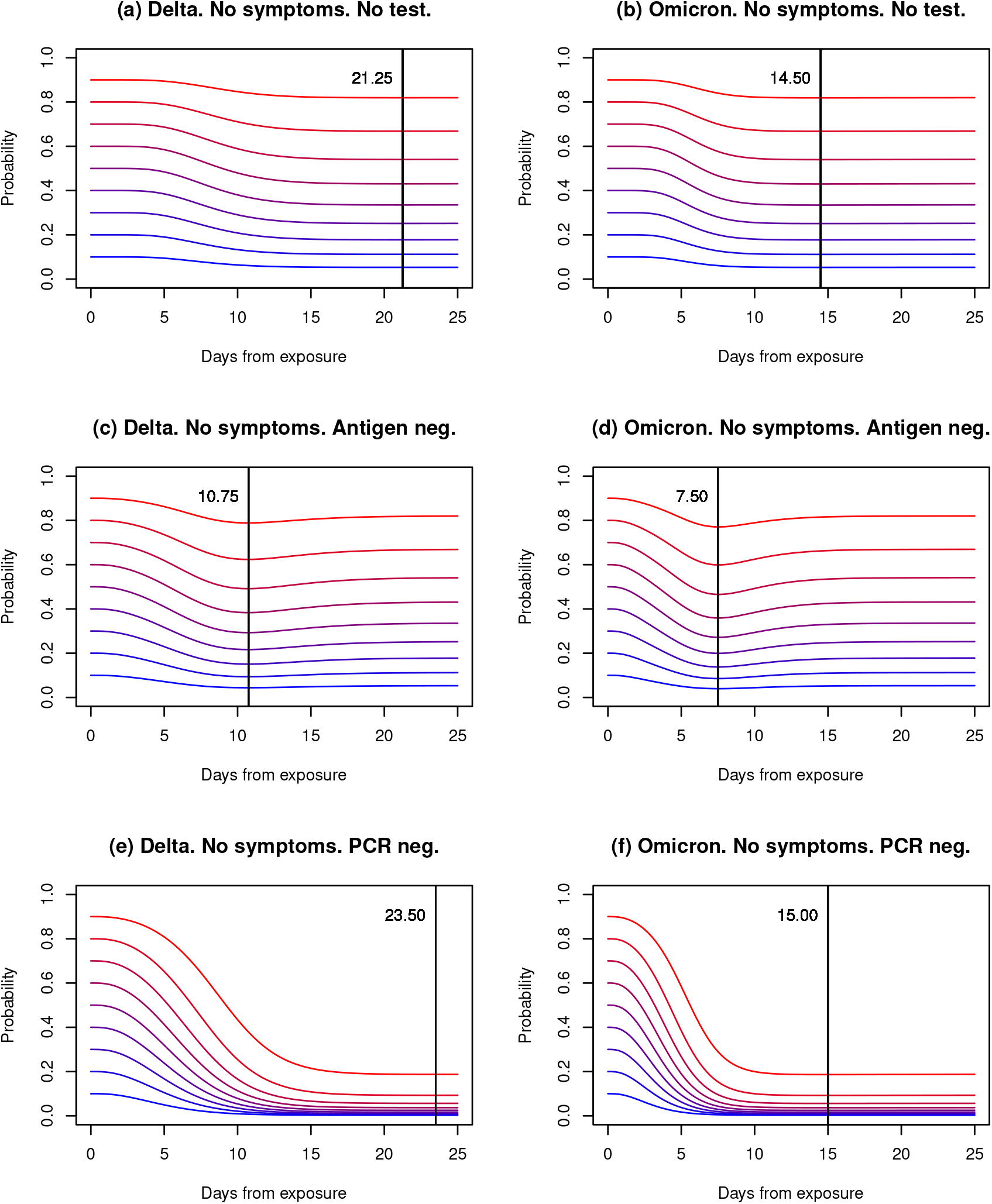
The posterior probability that an infection occurred at time 0 given a negative test following and no symptoms. The curves are given for varying prior probabilities. The vertical lines show where the posteriors are lowest.

## 4 Discussion

We have devised a software tool to assist decisions about the duration of quarantine and the timing of collection of diagnostic tests for individuals who have been exposed to a communicable disease. We provide examples of the use of the tool to support policies around quarantine for coronavirus disease 2019 infection caused by the Delta and Omicron SARSCoV-2 variants. The programs provide key insights about the relative utility of symptom monitoring alone versus symptom monitoring plus laboratory testing. Moreover, the software enables comparison of laboratory tests with different test characteristics. The tool allows the user to specify the assumed probability that an exposure resulted in transmission and set threshold probabilities for decisions about when to end quarantine. The tool makes it clear that evaluation of test results should not be done naively. A test may be negative because a transmission has not occurred, because the test result is false negative, because the individual has passed quickly through the course of the infection, or because the individual is experiencing an unusually long latent phase.

Figures 3 and 4 show how the state probabilities following exposure inform the interpretation of test results. Assuming a transmission risk of 30%, which is relatively high for most exposures, a negative antigen or PCR test, plus absence of symptoms, at about 7 days for Delta, and 5 days for Omicron denotes a 10% probability that an individual is infected. However, those that are infected are most likely in the latent phase and about to progress through the infection with risk to themselves and, potentially, others.

Figures 5 show that a strategy of repeat testing of a highly sensitive test does not shorten the time to reach a given infection threshold. In the example given, the time to reach the 1% threshold was virtually the same when one test was collected as when two tests were obtained, regardless of which tests were performed.

Figure 6 compared the extremes of definitely infected and definitely uninfected individuals. While waiting until the probability of infection curves cross is probably too conservative a criterion for ending isolation following exposure, it is important to remember that the risk from general contact is not zero. These results also show the significant benefit of antigen tests over PCR tests for detecting resolved cases. For instance, for the Omicron variant, a negative antigen test at 12 days indicates that there is a less than 10% chance that a symptom free individual is infected with the individual most likely having progressed to the recovered state. On the contrary, because the PCR test remains positive for the first portion of the recovered state, it has low utility for deciding on the duration of quarantine when the exposure certainly or almost certainly resulted in transmission. With a negative PCR test at 12 days, the probability of being infected, latent or infectious, was still about 25%, which is the same level as an untested individual whether symptom free or not. So, a PCR test at this time is worthless.

Figure 7 demonstrates the low predictive value of the negative antigen test for establishing that transmission did not occur in symptom-free individuals. Furthermore, we see a very narrow window in time, around 8 to 11 days for Delta, around 5 to 8 days for Omicron, for obtaining even this information with the curves reverting to the levels of the posterior probabilities given only that the individual was symptom-free. Conversely, the statistical power of PCR testing for confirming that a transmission did not occur is clearly seen in figures 7 (e) and (f), with clear consequences for case counting and population monitoring.

To some extent, many of the results presented above might have been anticipated because the different properties of PCR and antigen tests are well understood, however, even when the qualitative differences might be anticipated the quantitative results given here provide valuable insight that may be applied to creating public health or individualized policies for quarantine or isolation.

By differentiating the disease states and calculating their probabilities, it is possible to distinguish in a quantifiable way why a test result might be negative, namely, because there was no transmission, because the test is a false negative, because the individual has passed quickly through the course of the infection, or because they are experiencing an unusually long latent period. Since each of these reasons has different clinical implications, accurately assessing their likelihood is crucial. Importantly, by altering the input parameters, the tool can be extended to apply to other existing or future emerging communicable diseases that progress from latent to infectious to recovered states.

## Data Availability

Data sharing is not applicable to this article as no datasets were generated or analyzed during the current study.

## Availability and requirements

**Project name:** Covid isolation project.

**Project home page:** Not applicable.

**Operating systems**: Platform independent.

**Programming language:** `R`.

**Other requirements:** The `coga R` package.

**License:** GPL-2.

**Any restrictions to use by non-academics:** None.

## Supplemental files

`covid.R`: a text file containing `R` source code for the general programs for calculating disease state probabilities.

`covid.pdf`: a `pdf` file documenting the source code in `covid.R`. `isol.R`: a text file containing `R` source code for generating the figures presented in this article.

## Declarations

### Consent for publication

All authors have seen and approved the manuscript.

### Competing interests

The authors declare that they have no competing interests.

### Funding

ALH and MHS received funding from the Utah Office of Management and Budget for the Utah Health and Economic Recovery Outreach Project.

This work was supported by the Centers for Disease Control and Prevention Modeling Infectious Diseases in Healthcare, MInD-Healthcare, Program award U01CK000585.

This work was supported by Centers for Disease Control and Prevention (CDC) (SHEP-heRD 2021 Domain 1-A015, grant number 75D30121F00003).

## Notes

### Competing Interest Statement

The authors have declared no competing interest.

## References

Dinnes, J., Sharma, P., Berhanne, S., van Wyk, S. S., Nyaaba, N., Domen, J., Taylor, M., Cunningham, J., Davenport, C., Dittrich, S., Emperador, D., Hooft, L., Leeflang, M. M. G., McInnes, M. D. F., Spijker, R., Verbakel, J. Y., Takwoingi, Y., Taylor-Phillips, S., den Bruel, A. V., Deeks, J. J. & Cochrane COVID-19 Diagnostic Test Accuracy Group (2022), Rapid, point-of-care antigen tests for diagnosis of SARS-CoV-2 infection (review), Cochrane Database of Systematic Reviews.

Eikenberry, S. E., Mancuso, M., Iboi, E., Phan, T., Eikenberry, K., Kuang, Y., Kostelich, E. & Gumel, A. B. (2020), To mask or not to mask: Modeling the epotential for face mask use by the general public to curtail the COVID-19 pandemic, Infectious Disease Modelling 5, 293–308.

Hu, C., Pozdnyakov, V. & Yan, J. (2020), Density and distribution evaluation for convolution of independent gamma variables, Computational Statistics 35, 327–342.

Hu, C., Pozdnyakov, V. & Yan, J. (2021), coga: Convolution of Gamma Distributions. R package version 1.1.1.

Jansen, L., Tegomoh, B., Lange, K., Showalter, K., Figliomeni, J., Babdalhamid Iwen, P. C., Fauver, J., Buss, B. & Donahue, M. (2021), Investication of a SARS-CoV-2 B.1.1.529 (Omicron) variant cluster – Nebraska, November–December 2021, Morbidity and Mortality Weekly Report, CDC.

Liu, A. B., Davidi, D., Landsberg, H. E., Francesconi, M., Platt, J. T., Nguyen, G. T., Yune, S., Deckard, A., Puglin, J., adn D H Hamer, S. B. H. & Springer, M. (2022), Asociation of COVID-19 quarantine duration and postquarantine transmission risk in 4 university cohorts, JAMA Network Open 5, e220088.

Menni, C., Valdes, A. M., Polidori, L., Antonelli, M., Penamakuri, S., Nogal, A., Louca, P., May, A., Fiueirede, J. C., Hu, C., Molteni, E., Canas, L., Osterdahl, M. F., Modat, M., Sudre, C. H., Fox, B., Hammers, A., Wolf, J., Capdevila, J., Chan, A. T., David, S. P., Steves, C. J., Ourselin, S. & Spector, T. D. (2022), Symptom prevalence, duration, and risk of hospital admission in individuals infected with SARS-CoV-2 during periods of omicron and delta variant dominance: a prospective observational study from the ZOE COVID Study, Lancet pp. 1618–1624.

R Core Team (2015), R: A Language and Environment for Statistical Computing, R Foundation for Statistical Computing, Vienna, Austria.

Smith-Jeffcoat, S. E., Mellis, A. M., Grijava, C. G., Talbot, H. K., Schmitz, J., Lurrick, K., Ellingson, K. D., Stockwell, M. S., McLaren, S. H., Nguyen, H. Q., Rao, S., Asturias, E. J., Davis-Gardner, M. E., Suthar, M. S., Kirking, H. L. & RVTN-Sentinel Study Group (2024), SARC-CoV-2 viral shedding and rapit antigen test performace–Respiratory Virus Transmission Network, November 2022 - May 2023, Technical Report 16, CDC Morbidity and Mortality Weekly Report.

